# Impacts of patient advisory councils on recovery for sepsis survivors: a case study

**DOI:** 10.1101/2025.02.06.25321290

**Authors:** Mia Sheehan, Stefanie K Novakowski, Fatima Sheikh, Susan Korstad, Kristin MacDonald, Jordan Sacks, Kristine Russell, Marie-Maxime Bergeron, Marianne Vidler, Sepsis Canada

## Abstract

**Background:** Sepsis is a life-threatening condition with significant long-term impacts for survivors and their families. The known benefits of patient engagement have led to increased efforts globally to involve survivors in sepsis research. This study aimed to characterize the experiences of sepsis survivors and their families in patient advisory councils (PACs) for two Canadian sepsis research networks (Action on Sepsis and Sepsis Canada) and explore how PAC involvement supports long-term recovery.

**Methods:** This mixed-methods cross-sectional study consisted of a structured survey, ten interviews, and one focus group discussion. All current members of the Sepsis Canada and Action on Sepsis PACs (n=29) were invited to participate. The results of the survey were analyzed descriptively and used to inform the development of the semi-structured interview guide. Qualitative data were analyzed using a thematic approach.

**Results:** Overall, 15 PAC members participated. The majority of participants were women and over 40 years old. Survey scores showed that most participants felt meaningfully engaged, while the qualitative findings highlighted how PACs supported recovery and fostered community connections between survivors, families, and researchers. Major themes included sepsis experience, recovery journey, characteristics of PACs, characteristics of PAC participation, and impacts of PAC involvement.

**Interpretation:** Our findings demonstrate that PACs provide critical benefits that extend beyond feeling valued or appreciated for contributing to a specific project. These findings highlight the value of patient-oriented research in shaping evidence-based practices and policies and emphasize the need for trauma-informed approaches and improved post-sepsis care pathways to enhance recovery outcomes.

## Introduction

Sepsis is life-threatening organ dysfunction caused by a dysregulated host response to infection (1,2). In 2017, there were an estimated 48.9 million cases of sepsis and 11 million sepsis deaths globally (3). In Canada, a retrospective review of administrative data from Ontario found that 1 in 20 hospitalizations included sepsis, and healthcare costs for patients with sepsis were $672 million more than their matched controls (4). Patients who survive sepsis can experience long-term emotional, physical, neurocognitive, and economic impacts (5), including heightened risk of rehospitalization (6), increased use of high-cost healthcare (7), and increased risk of death (8,9). Caregivers and family members of individuals hospitalized for critical illnesses such as sepsis also experience ongoing psychological and emotional distress, including post-traumatic stress disorder (10). Given the significant impacts of sepsis on survivors and their families, there have been increased efforts to better incorporate patient and family voices in sepsis research.

Over the past decade, patient engagement has become a critical approach to enhance the relevance, quality, and impact of healthcare and research (11). The inclusion of lived experience allows for improved research and clinical care by ensuring that research questions, outcomes, and sharing of the results align with patient and community needs and priorities (12). This engagement can further contribute to patient empowerment, leading to transformative experiences for patient partners (13,14). Partnering with survivors and their families also enhances patient voices and contributes to patient-oriented research that informs policy (15) and health system change (16). Given the well-documented benefits of engaging with patients and caregivers to improve research and healthcare delivery (17), sepsis organizations globally (e.g., Health Quality BC, Canadian Sepsis Foundation, Global Sepsis Alliance, UK Sepsis Trust) have engaged with patients to raise awareness of sepsis and improve advocacy and clinical care through patient involvement. In the last five years, two Canadian sepsis research networks (University of British Columbia (UBC)’s Action on Sepsis Research Excellence Cluster and Sepsis Canada) established patient advisory councils (PACs) to guide strategic decision-making and facilitate patient-oriented research. However, there is limited literature characterizing research network engagement within the context of patients’ long-term recovery from sepsis, or other critical illnesses (e.g., post-ICU syndrome, long-COVID) (18). This study aimed to address this gap by exploring the experiences of sepsis survivors and their families participating in PACs and characterizing the impact of this engagement on their long-term recovery.

## Methods

### Study design

This mixed-methods cross-sectional study was initiated and led by two members of the Sepsis Canada and Action on Sepsis PACs and a UBC Assistant Professor (Appendix 1, Supplement 1). We referred to the Consensus-Based Checklist for Reporting of Survey Studies (19), Consolidated Criteria for Reporting Qualitative Research (20), and Guidance for Reporting Involvement of Patients and the Public (GRIPP2) (21) for reporting the study (Appendix 1, Supplement 2-4).

### Participant recruitment

All current members of the Sepsis Canada or Action on Sepsis PACs were eligible to participate. Members were either sepsis survivors or caregivers of sepsis survivors. Recruitment for the survey and interviews/focus group discussions were conducted separately, using targeted recruitment through the PACs membership email lists. Participants did not have to complete the survey to be eligible to participate in interviews.

### Data collection

The survey was administered using Research Electronic Data Capture and participants provided consent electronically. The survey (Appendix, Supplement 5) used the validated Patient Engagement in Research Scale (PEIRS) (21) and results were used to inform the development of the semi-structured interview guide (Appendix 1, Supplement 6). All interviews and focus group discussions were conducted virtually by MS and audio and video recorded on Zoom.

### Data analysis

Survey results were summarized using descriptive statistics. Interviews and the focus group discussion were transcribed verbatim and analyzed thematically (22) using NVivo 14 (QSR International, Melbourne, Australia). Our analysis was informed by a phenomenology approach, which focused on describing the lived experiences and understanding the meanings participants attributed to their experiences (23–25). Coding was done manually by MS and MV and all coded transcripts were reviewed by MMB, JS, and FS.

### Ethics statement

Ethics approval was obtained from the UBC/Children’s and Women’s Health Centre of British Columbia Research Ethics Board (H23-03622).

## Results

Overall, 15 individuals participated in the study (Figure 1). Most respondents were women, over 40 years old, and were sepsis survivors. Participants were from six provinces across Canada (ON, AB, BC, MB, SK, QC). Thirteen participants were only part of the Sepsis Canada PAC, 1 participant was only part of the Action on Sepsis PAC, and 3 participants were part of both PACs.

**Figure 1.**
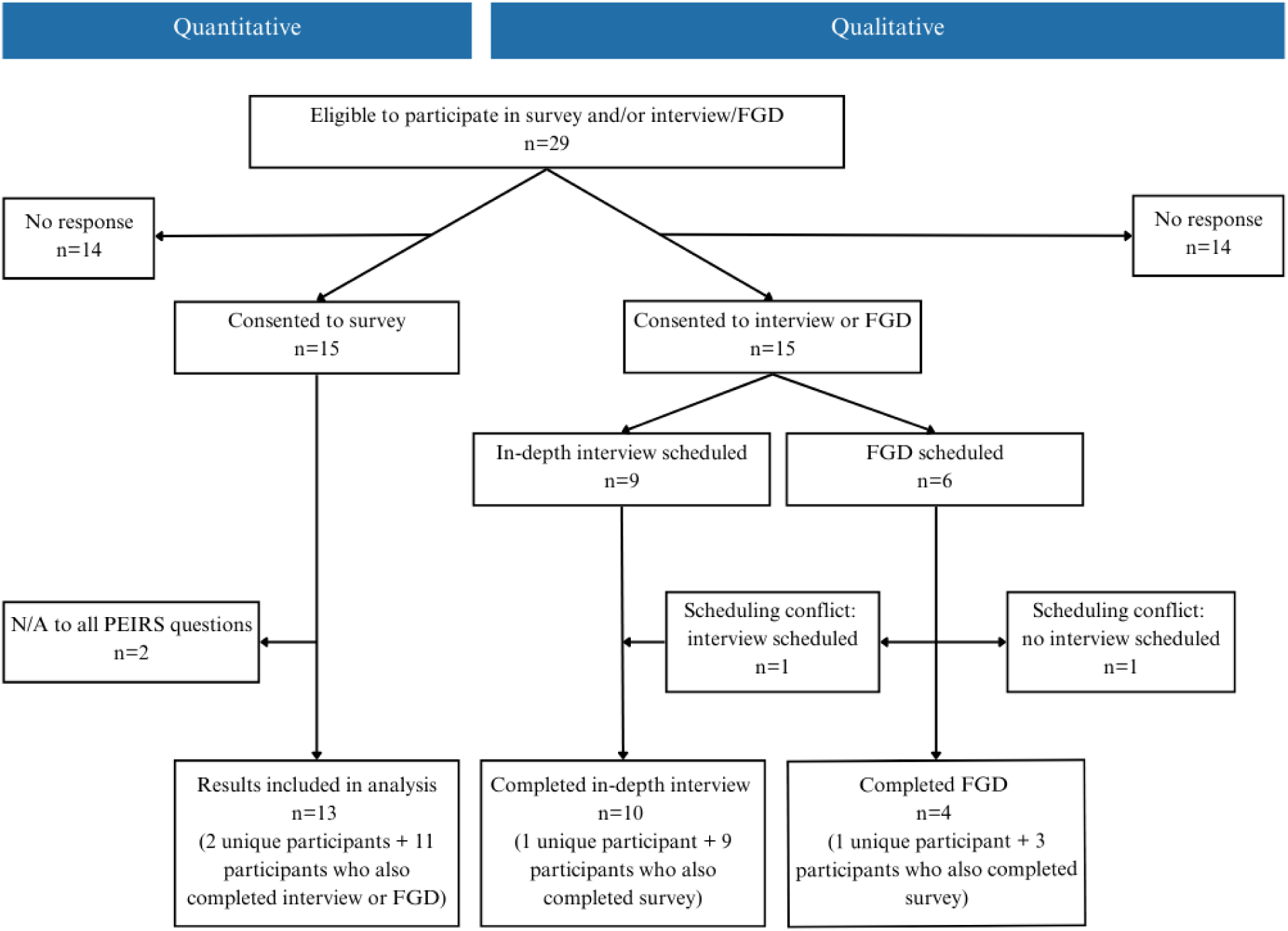
Participant flow chart for quantitative and qualitative methods. A total of 15 PAC members participated in the study.

Among the 13 participants who completed the survey, three participants reported an ‘extremely’ high degree of meaningful engagement, two reported a ‘very’ high degree, seven reported a ‘moderate’ degree, and one participant reported a ‘deficient’ degree of meaningful engagement (Table 1). Across individual domains and participants, we observed scores consistent with a deficient degree of meaningful engagement in convenience, team environment and interaction, support, and benefits. We focused the interview questions on these topics.

**Table 1.**
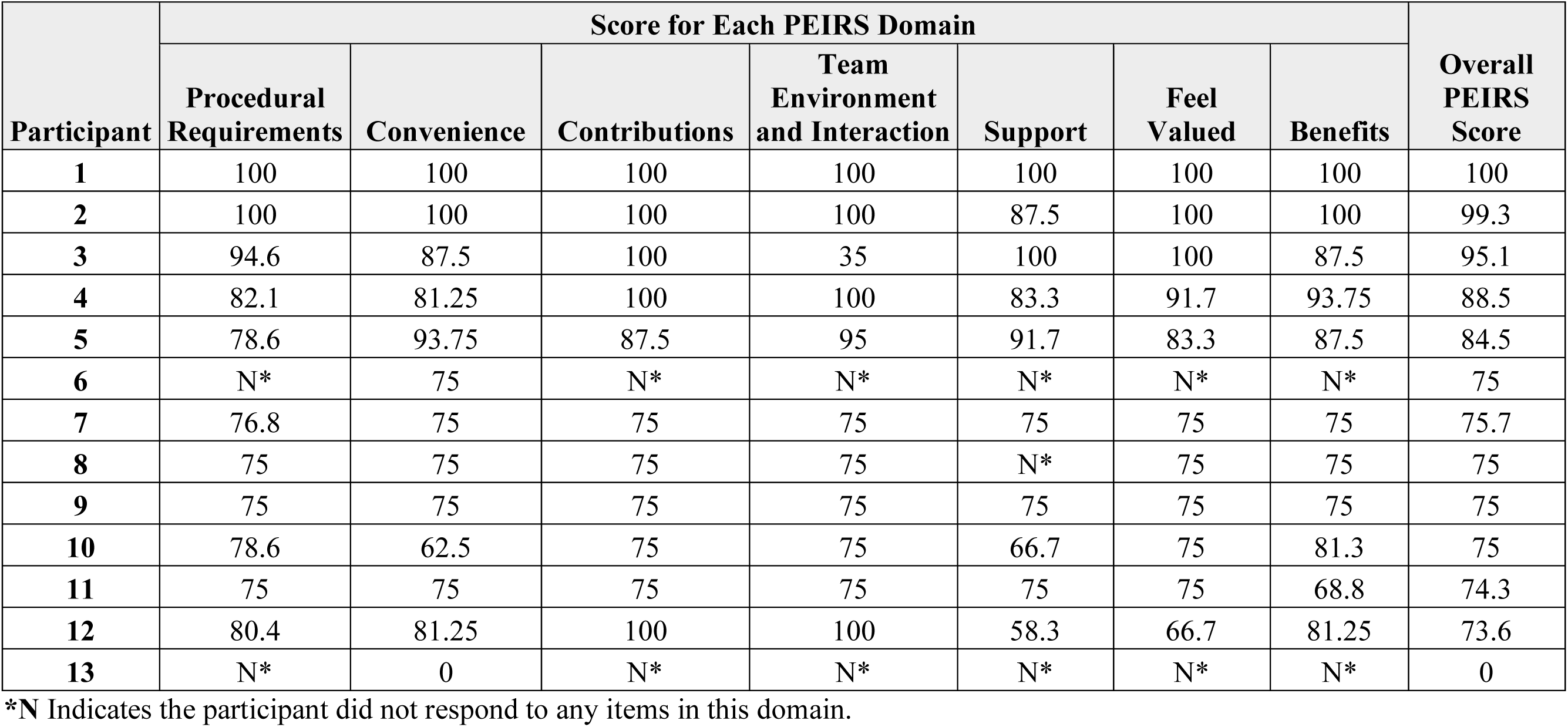
Level of meaningful patient engagement across the PEIRS domains (n=13). In the validated PIERS (20), a score below 70.1 is considered ‘deficient’ degree of meaningful engagement; between 70.1 and 82.7 is a ‘moderate’ degree; between 82.7 and 92.0 is a ‘very high degree’; and above 92.0 is an ‘extremely’ high degree. The maximum score possible is 100.

| Participant | Score for Each PEIRS Domain |  |  |  |  |  |  | Overall PEIRS Score |
| --- | --- | --- | --- | --- | --- | --- | --- | --- |
|  | Procedural Requirements | Convenience | Contributions | Team Environment and Interaction | Support | Feel Valued | Benefits |  |
| 1 | 100 | 100 | 100 | 100 | 100 | 100 | 100 | 100 |
| 2 | 100 | 100 | 100 | 100 | 87.5 | 100 | 100 | 99.3 |
| 3 | 94.6 | 87.5 | 100 | 35 | 100 | 100 | 87.5 | 95.1 |
| 4 | 82.1 | 81.25 | 100 | 100 | 83.3 | 91.7 | 93.75 | 88.5 |
| 5 | 78.6 | 93.75 | 87.5 | 95 | 91.7 | 83.3 | 87.5 | 84.5 |
| 6 | N* | 75 | N* | N* | N* | N* | N* | 75 |
| 7 | 76.8 | 75 | 75 | 75 | 75 | 75 | 75 | 75.7 |
| 8 | 75 | 75 | 75 | 75 | N* | 75 | 75 | 75 |
| 9 | 75 | 75 | 75 | 75 | 75 | 75 | 75 | 75 |
| 10 | 78.6 | 62.5 | 75 | 75 | 66.7 | 75 | 81.3 | 75 |
| 11 | 75 | 75 | 75 | 75 | 75 | 75 | 68.8 | 74.3 |
| 12 | 80.4 | 81.25 | 100 | 100 | 58.3 | 66.7 | 81.25 | 73.6 |
| 13 | N* | 0 | N* | N* | N* | N* | N* | 0 |

\*N Indicates the participant did not respond to any items in this domain.

Information shared by PAC members was characterized into five major themes: sepsis experience, recovery journey, PAC characteristics, characteristics of PAC participation, and impacts of PAC involvement. Sub-themes and illustrative quotes are available in Tables 2–6.

**Table 2.** Sub-themes and illustrative quotes related to the sepsis experience.

| Theme | Key findings | Supporting quote |
| --- | --- | --- |
| **Sepsis experience** | • Diverse experiences with sepsis<br>• Highlighted importance of timely and accurate care<br>• Majority were in medically induced comas in the ICU, though one participant was unable to be admitted due to the shortage of available beds<br>• Many participants were never informed they had sepsis | *“I felt like I wasn’t really relayed the proper information that I should have had as a caretaker. And for me, I lost a bit of hope in... the healthcare system.” - P11 (caregiver)* |

### Sepsis experience

Regardless of the cause, participants’ sepsis experiences were sudden, severe, and life-altering (Table 2). Many participants faced misdiagnosis and late detection, resulting in severe cases of sepsis. Others experienced sepsis as an unexpected complication following surgery. Both survivors and their families frequently expressed receiving poor communication from healthcare providers regarding sepsis diagnosis and post-discharge care.

### Recovery journey

Most participants had no expectations for recovery, due to the lack of information shared at discharge about the mental, physical, and cognitive impacts of sepsis that they would experience. Some survivors were never told that they had sepsis, contributing to their lack of expectations. Participants described many physical impacts (Table 3) that affected their ability to complete everyday activities. All participants faced significant mental health challenges following sepsis, and some reported post-traumatic stress disorder and suicidality. Survivors often felt isolated and alone in their care and recovery. Many survivors sought psychiatric care for mental health support, often on their own, as this support was not integrated into their sepsis care. Cognitive challenges were described as a common result of sepsis by the participants. These difficulties, including memory loss, impacted their daily functioning and ability to return to work or engage in certain PAC activities. Caregivers and other family members struggled witnessing their loved ones experience sepsis and navigating decision-making, in part due to the lack of information and support provided for caregivers and family members.

**Table 3.** Sub-themes and illustrative quotes related to characteristics of recovery journey.

| Sub-themes | Key Findings | Illustrative quote |
| --- | --- | --- |
| Recovery expectations | • Had no expectations of the length of recovery and when everyday activities could be resumed, including returning to work<br>• Caregivers were unaware of long-term impacts of sepsis and how to better support their family member | *“I didn’t know I was a sepsis survivor. Those words [were] actually never used with me in the hospital. So, when I was discharged, I had no expectations. I didn’t know what had been wrong with me.” - P4 (sepsis survivor)* |
| Physical recovery | • Common physical symptoms among survivors following discharge: low energy, difficulty walking and completing other everyday activities, weight loss, and insomnia<br>• Other long-term physical impacts of sepsis and its complications: chronic pain, nerve damage, amputations, and bowel resections | *“[Sepsis] drastically impacted [my physical health] and I’m permanently disabled.” - P5 (sepsis survivor)*<br>*“Because I... was on a ventilator, I couldn’t speak. I had to relearn how to speak and breathe [on my own].” - P8 (sepsis survivor)* |
| Mental recovery | • Processing trauma was particularly difficult due to delayed awareness caused by being in a medically induced coma, resulting in survivors having to cope with events that had already occurred<br>• Faced uncertainties of how long their recovery would take or whether they | *The fact that no [healthcare provider] was kind of looking out for me despite me raising concerns, and my husband raising concerns... truly made me realize that... patients are out there on their own.” - P10 (sepsis survivor)* |
|  | could ever fully return to their pre-sepsis lives<br>• COVID-19 pandemic was a trigger for many sepsis survivors and brought traumatic memories, due to media coverage of ICU shortages and the fear of contracting COVID-19, which can lead to sepsis<br>• Mental health challenges were translated into motivation for becoming involved in PACs | *"I was very emotional, I was angry, I was sad... I was really lonely... No one knew what I'd been through... So it was a tough time."* - P4 (sepsis survivor) |
| Cognitive recovery | • Cognitive difficulties included: forgetfulness, memory loss, and false memories of being in the ICU<br>• Challenges highlighted need for better reminders about meetings and roles with the council | *"Part of my sepsis journey has been... having a lot of memory issues... a lot of focus issues, [and] executive functioning issues."* - P5 (sepsis survivor) |
| Economic recovery | • Difficulties meeting the physical and cognitive demands of their job after returning to work following their sepsis experience<br>• Many participants were only able to return to work part-time, while some were able to resume full-time work<br>• A few participants were on long-term disability due to the lasting effects of sepsis | *"I tried to go back to work at the same place. Just because of my... strength and determination, no one was going to tell me I [couldn't] go back to work. So, I went back twice, but it ended up being too difficult. The obstacles in terms of... the number of hours that I had to work and the physical demand of the job... It's not a physically demanding job in itself, but everything for me is... work now."* - P12 (sepsis survivor) |
| Others' recovery | • Survivors recognized the impacts that sepsis had on their family members and difficulties they faced in their recovery | *"The whole sepsis recovery takes a lot out of you as a patient, but even more so as a family member."* - P14 (sepsis survivor) |

### PAC characteristics

PACs were composed of empathetic and supportive individuals with a common goal of advocating for improvement in sepsis care and research (Table 4). Many participants highlighted the need to engage with more family members and caregivers, as *“they’ve had different lived experiences*” (P6 – sepsis survivor) that could enrich the work of the councils. Some respondents felt the council would benefit from a wider range of values and insights, as they felt certain voices were dominating the decision-making processes. Most participants valued the recognition and compensation that they received through their PAC involvement.

**Table 4.** Sub-themes and illustrative quotes related to PAC characteristics.

| Sub-themes | Key Findings | Illustrative Quote |
| --- | --- | --- |
| Council composition | • Majority of PAC members were sepsis survivors | *"I meet a lot more patient partners that have actually* |
|  | • Participants understood importance of having more diversity in PACs<br>• Highlighted the lack of male representation in PACs | *had sepsis themselves... Whereas... I haven't met very many people that... are similar to me, where they were actually caregivers" - P11 (caregiver)* |
| Recognition and compensation | • Recognition included being acknowledged as patient partners in grant applications, manuscripts, other knowledge translation materials, and receiving invitations to speak at conferences<br>• Financial compensation was not necessary but was viewed as a meaningful gesture of appreciation<br>• Participants required a primary source of income beyond their role within the PAC, which reduced the time they had available to contribute to the councils | Respondents discussed verbal and written appreciation they received, and feeling like their contributions were valued, particularly when they could *"see... the results of [their] contributions in a tangible way"*. - P14 (sepsis survivor) |
| Meeting accommodation and format | • It was acceptable for participants to miss meetings if a work or personnel conflict arose, and meetings notes were made accessible afterwards<br>• Meetings consisted of opportunities for members to participate and have discussion | *"I love that it's online... I love that we can have people from... [the] east coast to the west coast participate... within their own time zones. We do try to have a lot of... accessibility features, so people can connect online, they don't have to have their camera on, they don't have to talk... they can just be... quiet, passive individuals... We do ask for people's contributions and people are able... to talk during our sessions. They can... send an email afterwards... There [are] other ways to... submit their... thoughts and ideas."* - P5 (sepsis survivor) |
| Communication | • Clear understanding among participants that the councils welcomed feedback and questions during meetings | *"I think there's really good communication with the council, but to an amount where it's not* |
|  | Participants felt engaged, informed, and up to date on study opportunities and outcomes of council activities | *overwhelming.” - P15 (sepsis survivor)* |
| Activities | Activities included participating in meetings and research projects, conducting presentations, reviewing grant applications, and informing the development of a National Sepsis Action Plan<br>Many of the projects were tailored towards the interests, skill sets, and experiences of the participants<br>Members’ roles included patient partner, steering committee member, and co-chair of PAC<br>Participants described the time commitment as appropriate and manageable | Many participants were also engaged in related committees and councils, as their involvement in sepsis PACs “*opened up some doors to... speak to other people and get involved in other groups.*” - P11 (caregiver) |

Responses from the participants agreed that PAC meeting times and outlines, study opportunities, and commitments were well communicated. PAC members were often consulted regarding availability, and all meetings were conducted virtually. However, those who were able to join PAC meetings during work hours emphasized the challenge of making a *‘mental switch’* (P8 – sepsis survivor), switching quickly between their roles as a sepsis survivor and their professional roles beyond the PAC. Further, some respondents noted the lack of clarity regarding roles and responsibilities in the council, and the need for more frequent reminders of the committee’s purpose and structure. Participants often struggled to characterize the specific activities that they contributed to, and did not always distinguish between activities led by the PACs, research networks, or individual research teams.

### Characteristics of PAC participation

Respondents initially had no expectations regarding their participation in PACs, largely due to the lack of specificity in communications describing how members could contribute to the PACs and the impacts of this involvement (Table 5). Many participants were surprised, stating they *“didn’t expect for [their] voice to be so valued”* (P2 – sepsis survivor). A common reason for joining PACs was to better understand what had happened to them or their family members during their experience with sepsis. Using this knowledge, participants wanted to advocate for sepsis awareness among the public and healthcare workers to ensure earlier diagnoses for future sepsis patients and improve communication about post-discharge support to enhance the recovery of current and future sepsis survivors. Another key motivator for joining PACs was to connect with other sepsis survivors and caregivers.

**Table 5.**
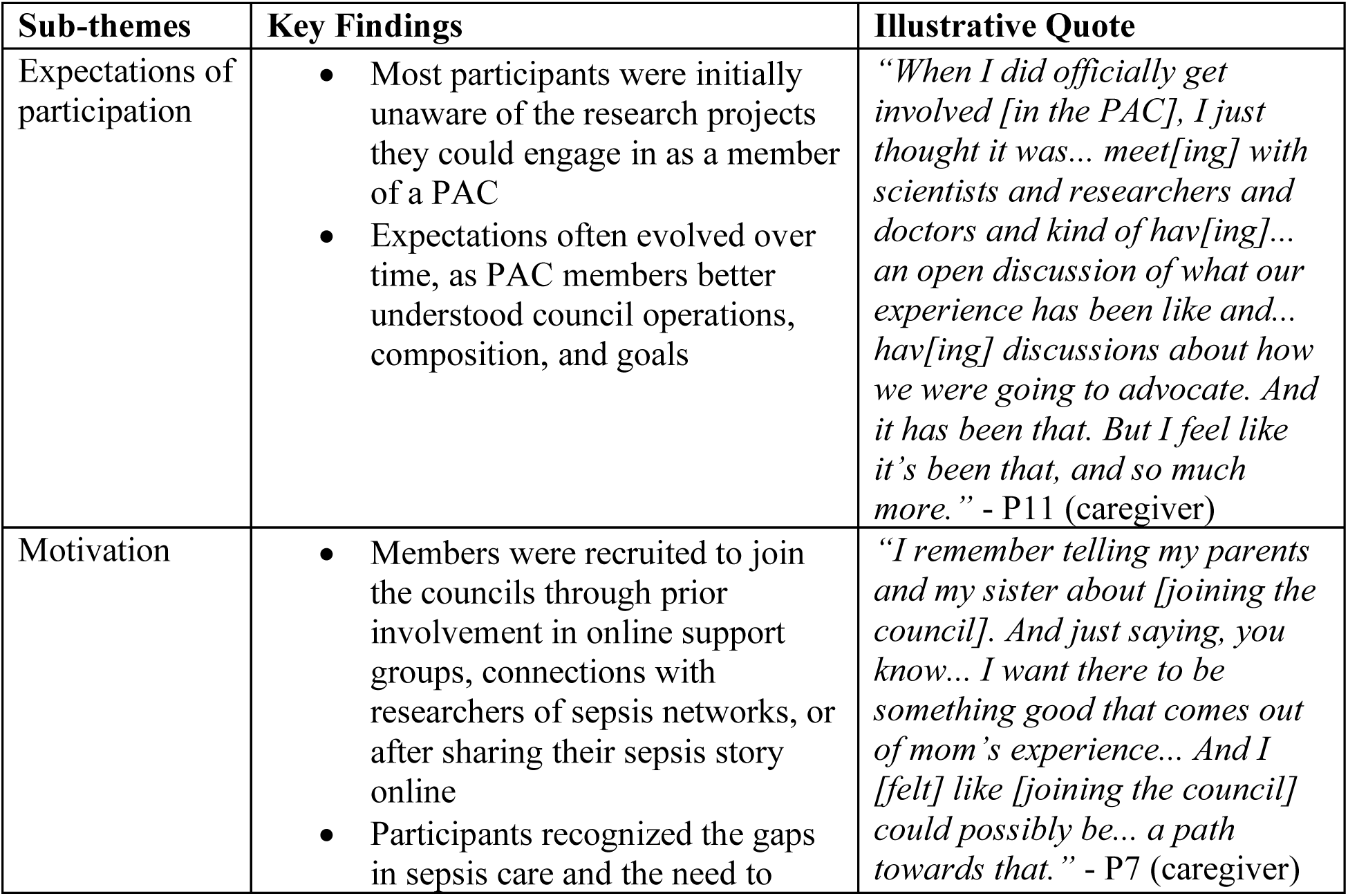
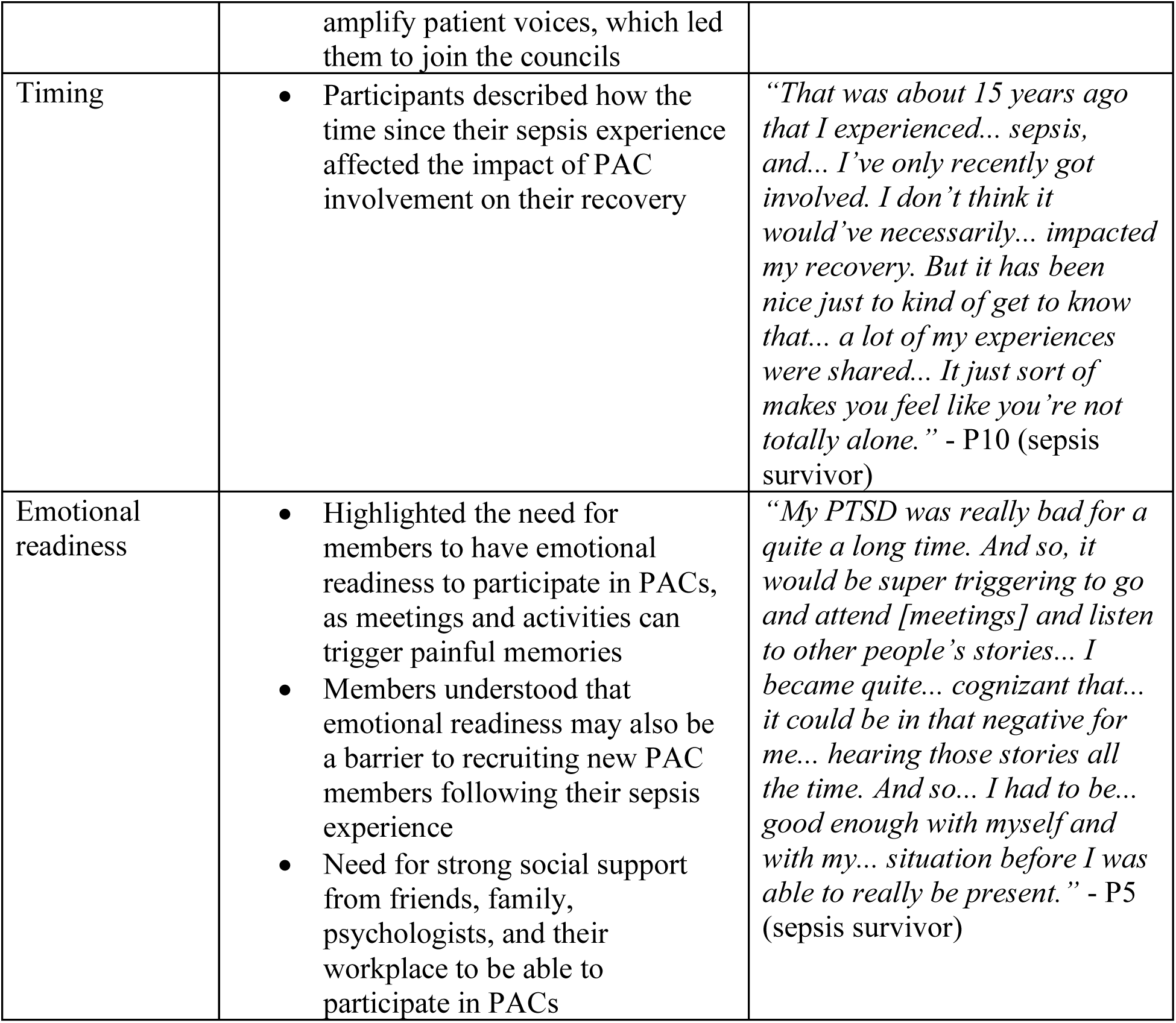
Sub-themes and illustrative quotes related to characteristics of PAC participation.

| Sub-themes | Key Findings | Illustrative Quote |
| --- | --- | --- |
| Expectations of participation | Most participants were initially unaware of the research projects they could engage in as a member of a PAC<br>Expectations often evolved over time, as PAC members better understood council operations, composition, and goals | *“When I did officially get involved [in the PAC], I just thought it was... meet[ing] with scientists and researchers and doctors and kind of hav[ing]... an open discussion of what our experience has been like and... hav[ing] discussions about how we were going to advocate. And it has been that. But I feel like it’s been that, and so much more.” - P11 (caregiver)* |
| Motivation | Members were recruited to join the councils through prior involvement in online support groups, connections with researchers of sepsis networks, or after sharing their sepsis story online<br>Participants recognized the gaps in sepsis care and the need to | *“I remember telling my parents and my sister about [joining the council]. And just saying, you know... I want there to be something good that comes out of mom’s experience... And I [felt] like [joining the council] could possibly be... a path towards that.” - P7 (caregiver)* |
|  | amplify patient voices, which led them to join the councils |  |
| Timing | Participants described how the time since their sepsis experience affected the impact of PAC involvement on their recovery | *“That was about 15 years ago that I experienced... sepsis, and... I’ve only recently got involved. I don’t think it would’ve necessarily... impacted my recovery. But it has been nice just to kind of get to know that... a lot of my experiences were shared... It just sort of makes you feel like you’re not totally alone.” - P10 (sepsis survivor)* |
| Emotional readiness | Highlighted the need for members to have emotional readiness to participate in PACs, as meetings and activities can trigger painful memories<br>Members understood that emotional readiness may also be a barrier to recruiting new PAC members following their sepsis experience<br>Need for strong social support from friends, family, psychologists, and their workplace to be able to participate in PACs | *“My PTSD was really bad for a quite a long time. And so, it would be super triggering to go and attend [meetings] and listen to other people’s stories... I became quite... cognizant that... it could be in that negative for me... hearing those stories all the time. And so... I had to be... good enough with myself and with my... situation before I was able to really be present.” - P5 (sepsis survivor)* |

Participants joined PACs at various stages of their sepsis recovery, ranging from a few months to over 10 years after their initial sepsis experience. Those who joined years after their sepsis experience reported less direct impact on their sepsis recovery. However, these participants recognized other positive effects, such as the normalization of their experiences and a strengthened sense of community. Participants emphasized the importance of understanding one’s mental and physical capacities and ensuring that involvement has a *“net benefit”* (P5 – sepsis survivor) on their recovery from sepsis.

### Impacts of PAC involvement

All participants mentioned that PAC involvement can impact community sepsis awareness, recognition, management, and recovery (Table 6). Participants highlighted some of the impacts that their PAC involvement has on the council itself, including benefits brought to the council from applying their professional skills and expertise to council activities and how they contributed to growing council membership by raising awareness of the impacts of PAC involvement. PAC members were able to better understand sepsis and research as a result of their involvement in the councils and were able to share the knowledge with family members which further *“helped them in their [mental and emotional] recovery*” (P11 – caregiver). Engaging in PACs allowed members to share their sepsis experience(s) with others, leading to feelings of fulfillment, strength, and connection with fellow survivors and their families.

**Table 6.** Sub-themes and illustrative quotes related to impacts of PAC involvement.

| Sub-themes | Key Findings | Illustrative Quote |
| --- | --- | --- |
| Impact on community | Connected with other sepsis survivors through online groups or posts on an individual level and provided one-on-one support with their recovery<br>Directly witnessed the impacts of their involvement on their community (e.g. raising awareness of sepsis and the importance of early recognition and diagnosis among their colleagues working in healthcare) | *“After being on [the council] for the last few years, I really discovered how much of an impact it makes having... patient partners being apart [of councils] and really advocating. Because they’re the ones that have actually experienced it. And it can* |
|  |  | *actually speak towards... the general public but also... researchers or healthcare workers.” - P11 (caregiver)* |
| Impact on council | • Noted the importance of further recruitment to improve and strengthen the council<br>• The impact of their PAC involvement on other council members depended on whether participants felt they brought unique perspectives or skills, their connections to other members, and whether they were comfortable sharing their stories regularly | *“There’s got to be some others that come in and can offer some other perspectives, so that we’re not hearing the same perspective all the time... And that might... breed some further ideas into where we need to be.” - P9 (sepsis survivor)* |
| Impact on immediate network | • PAC involvement had indirect positive impacts on family members of sepsis survivors<br>• However, some sepsis survivors mentioned that their family did not want to be involved in PACs as part of their recovery journey | *“If [my family] see that I’m doing very well and happy and it helped me process... my own experience, they would benefit from that as well.” - P1 (sepsis survivor)* |
| Impact on oneself | • Participants’ repeated exposure to discussions about sepsis and survivor experiences, along with sharing their stories, contributed to their mental healing<br>• Engagement in sepsis research and advocacy strengthened their knowledge of sepsis and understanding of their own experiences, aiding them in their recovery journey<br>• PAC opportunities also led to new connections with researchers and healthcare providers | *“Everyone’s situation is very unique... But at the end of the day, we know what the impact of sepsis and the toll it has. So, I think there still is that unity despite having very different experiences and stories to tell.” - P11 (caregiver)* |

### Interpretation

We explored the impacts of engaging with PACs on long-term recovery of sepsis survivors and their families. In our study, participants’ experiences with sepsis and recovery were validated through social interactions and improved knowledge of sepsis, creating positive impacts beyond feeling valued for their contributions through recognition or compensation. These findings add to prior research demonstrating the benefits of patient-oriented research (26, 27). We also found that members had limited expectations about their role and the impact of engaging in PACs and highlighted the ongoing challenge of establishing PACs that are truly representative of all patient populations.

Our study demonstrated that PAC involvement supports recovery, particularly the mental well-being of sepsis survivors and their families. Patient partners reported being able to build professional and personal communities and connections with other individuals with lived experience and build knowledge and skills surrounding sepsis. These positive impacts on patient well-being align with a recent study that found patient partners involved in a health research network for chronic pain used their involvement to build a social network that supported recovery and learning. This helped partners cope with their own pain experiences (26). In our study, patient partners also noted how sharing stories and experiences positively impacted their mental healing and recovery, similar to the experiences reported by ICU survivors participating in peer support groups (28).

We found that participants often had no expectations about their involvement in PACs when first joining the council. As research progressed, patient partners better understood the ways of participating in research, specific project goals, and team member needs, leading to clearer expectations for engagement. Similar experiences were reported by members of PAC for a kidney health research network (27). Within the context of health research networks, PAC members may support several research studies and knowledge translation activities with different aims, levels of engagement, and intended impacts (29). This breadth of activities may necessitate clearer communication about the roles and responsibilities of patient partners, research staff, and investigators, and the differences between individual-, network-, and council-led activities.

### Future Recommendations

The in-hospital sepsis experiences and post-discharge challenges described in our study re-enforce the need for enhanced patient-healthcare provider communication on the long-lasting impacts of sepsis and improved post-sepsis care pathways in Canada, and globally. There have been efforts to improve the hospital to home transition for sepsis patients (30), but these practices have not been widely implemented. Our study highlights the important role that patients and families should have as research partners involved in developing and evaluating post-sepsis care pathways and as individuals who support their peers in recovery as part of these pathways. Improved communication about roles and the benefits of engagement may enable recruitment of more diverse voices of sepsis survivors and their families as research partners. Addressing this gap in current research is critical, particularly in sepsis research, as equity-deserving communities face a disproportionately high burden of sepsis (3, 31, 32). Educational materials on post-sepsis syndrome provided to sepsis survivors at discharge may be a venue for communicating benefits and recruiting additional partners; however, careful consideration of the timing of recruitment relative to a patient’s journey and processes for supporting safe and respectful engagement is needed.

### Limitations

Due to our sampling population, which only included current PAC members, our results are biased towards positive experiences with PACs and recovery from sepsis. Further, our results may not be generalizable given the small sample size and the limited sociodemographic diversity within our study population. The small sample size also made it difficult to assess the impacts of specific PAC activities or characteristics on recovery. Participants also experienced difficulty distinguishing between specific activities, further contributing to this limitation and highlighting the need for clearer communication by the research networks. Despite these limitations, the findings are strengthened by the study team’s extensive experience in sepsis research and their lived experiences with sepsis, bringing a unique and comprehensive perspective to the study.

## Conclusion

Our study demonstrates that PACs provide benefits go beyond participants’ sense of contributing to a specific project and support recovery. Insights from this study reinforce the value of patient-centered sepsis care and emphasize the importance of patient-oriented research in shaping evidence-based practices and policies. Through such engagement, Canada has an opportunity to develop inclusive and comprehensive sepsis care frameworks that better meet the needs of survivors and their families and reduce the long-term impacts of sepsis. A national sepsis strategy, as called for by the World Health Assembly 70.7 resolution on sepsis (33), would facilitate coordinated implementation of such frameworks (34). Notably, this study is among the first to explore how engagement with research networks supports recovery for sepsis survivors, offering a model for similar initiatives and highlighting the potential for research networks to drive meaningful improvements in patient recovery and care.

## Contributions

The contributions of the authors of the manuscript are as follows: SN, FS, KM, SK, KR, MMB, and MV contributed to conceptualization and design. MS and SN collected data. MS, SN, JS, MMB and MV analyzed data. MS drafted the manuscript and SN, FS, and MV edited the manuscript. All authors contributed to interpreting data and reviewing the manuscript. All authors approved the final version to be published and agreed to be accountable for its accuracy and integrity.

## Data Availability Statement

The datasets generated and analyzed during the current study are not publicly available due to their potentially identifiable nature. They are available from the corresponding author on reasonable request.

