## Appendix 1 for "Impacts of patient advisory councils on recovery for sepsis survivors: a case study"

### Supplement 1: Additional Details on Study Methods

*Study design and population*

This explanatory sequential mixed-methods study invited all current members of Action on Sepsis and Sepsis Canada Patient Advisory Councils (PACs) to participate. The UBC's Action on Sepsis PAC was established in May 2020 and consisted of 5 patient partners from three provinces. The Action on Sepsis PAC supports a provincial sepsis research network where most members are UBC faculty. The Sepsis Canada PAC was established in January 2022 and consisted of 28 patient partners across 8 provinces, including individuals who are also members of the Action on Sepsis PAC. The Sepsis Canada PAC supports a national sepsis research network with members from 8 Canadian provinces and 11 additional countries.

*Study team*

The study team consisted of 5 patient partners (MMB, KR, KM, SK, JS), a UBC Assistant Professor with experience in qualitative methodology and patient-oriented research (MV), the Action on Sepsis Network Coordinator (SN; PhD-prepared UBC employee who has supported Action on Sepsis and its PAC for the past 5 years), a PhD candidate and health equity specialist with experience conducting sepsis-related research in Canada (FS; trainee with Sepsis Canada), and a research assistant with Action on Sepsis (MS; recent MPH graduate who received training in conducting qualitative interviews and applying a trauma-informed lens to understanding patient experiences by MV).

*Participant recruitment*

All current members of the Sepsis Canada or Action on Sepsis PACs were invited to participate (n=29). Study invitation emails contained a short description of the study, contact information, and a link to the survey or contact information for the research assistant facilitating interviews and focus group discussions. These invitations were distributed by the Sepsis Canada or Action on Sepsis Network Coordinators after review and approval by the respective PAC Co-Chairs. After two weeks, a second email reminding members about the study was also distributed.

We recruited participants for the survey from February to March 2024, until the target sample size was reached (n=15). The sample size assumed a 50% response date among the 29 potential participants. The survey link remained active for one month after the recruitment period. After completing the survey, participants were automatically re-directed to a new Research Electronic Data Capture (REDCap) survey where they were invited to provide their name (optional) and email to receive a near cash equivalent (gift card) of $10 in recognition of their contribution. This enabled identifiers (name and email) to be stored separately from survey responses to ensure confidentiality of responses.

We recruited participants and conducted qualitative interviews and focus group discussions between May and August 2024, until data saturation was reached. PAC members who indicated their interest in participating in the interview were first contacted by a female research assistant

(MS; MPH) to schedule an initial review of the consent form, including the study objectives, overview of interview, and preference for focus group discussion or interview.  MS had no prior relationship with the PACs or sepsis research networks prior to this contact. During the initial review, MS introduced herself and her role in the study. Participants were provided with the option of either participating in interviews or focus group discussions as we recognized that some participants would receive emotional support from discussing these issues with peers who have similar experiences, while others prefer one-on-one interviews. Participants were given one week to review the consent form and make any inquiries to the study team before providing an electronic copy of their signed informed consent to MS. MS then scheduled either an interview or focus group discussion. The focus group discussion was scheduled once an appropriate group size (n=6) was reached.

Participants were offered $25 per hour (in the form of a gift card) for their time participating in the in-depth interview or focus group discussion. These individuals were reimbursed for additional costs (i.e., childcare) incurred as a result of participating in this study if discussed with MS before the interview took place. Participants were informed that they would still receive compensation even if they needed to end the interview early.

*Data collection – survey*

The survey was administered using REDCap hosted at the British Columbia Children's Hospital Research Institute. The Patient Engagement in Research Survey (PEIRS) (21) was selected based on relevance and appropriateness of the survey items to the study objectives. Prior to beginning data collection, additional questions on patient demographics and length and level of engagement with PACs were developed and iteratively revised following discussions among the study team. We pilot tested the survey for clarity and appropriateness with two PAC members. No significant changes were made to survey questions after pilot testing and responses from these two patient partners were included in the final analysis. No actions were taken to prevent multiple participation, as we determined this was unlikely given our targeted approach to recruitment.

*Data collection – interviews and focus groups*

A semi-structured interview guide was developed by the study team (SN, MV, MMB, KR, FS). The interview guide was pilot tested with a member of the study team who was not involved in creating the interview guide. No significant changes were made to the interview guide and the response from the patient partner was included in the final analysis. Interview topics included: sepsis experience, recovery journey, impact of PAC engagement, and experience with PACs. A trauma-informed lens was used to minimize potential triggering of emotional stress related to sepsis experiences while enabling open-ended exploration of the experiences of sepsis survivors and their families. Prior to beginning the interviews, all participants were provided with a list of appropriate, available, and affordable mental health services and reminded they could stop the interview at any time.

The interviews were 30 to 60 minutes, and the focus group discussion was 90 minutes. All interviews and the focus group discussion were conducted on Zoom from a private location within the research team's offices and participants were free to join from a location of their choice. No repeat interviews were conducted. MV attended the focus group discussion and first interview conducted to provide supervisory support for MS. MV had previously met two participant (virtually), as these individuals were also members of the study team, but otherwise had no prior relationship with the participants. MS, MV, and SN met after every interview to discuss field notes and progress to data saturation. One participant dropped out after consenting to a focus group discussion due to scheduling conflict, one participant initially scheduled for a focus group discussion completed an interview, and no interviews ended early. No interview transcripts were returned to participants. However, a summary of results was prepared and distributed to participants after data analysis was complete.

*Data analysis – survey*

Given the small sample size, the risk of re-identification is high and thus only aggregate data about patient demographics and engagement with PACs are reported. PEIRS scores were calculated following the published method (21). Each item on the PEIRS was assigned a numeric value between 0 and 4 (strongly disagree to strongly agree). Scores for each domain were calculated as: ((sum of responses)/(total number of items responded to within that domain x 4) *100). If participants responded as 'Not Applicable', no numerical value was assigned to that item and the denominator was modified accordingly. In the validated PIERS (21), a total score below 70.1 is considered a 'deficient' degree of meaningful engagement; between 70.1 and 82.7 is a 'moderate' degree; between 82.7 and 92.0 is 'very high' degree; and above 92.0 is an 'extremely high' agree.

*Data analysis – interviews and focus groups*

Inductive thematic coding was used to allow themes to emerge from the qualitative data (25) and for the perspectives of sepsis survivors and their families to be highlighted. The transcripts of three interviews were reviewed by the research team (MS, MV, and SN) who identified key themes and developed an initial coding framework. These researchers consistently discussed the frames and their interpretation of the qualitative data with a patient partner (MMB) until all coding frames were agreed. Coded transcripts were de-identified prior to sharing with additional team members to protect the confidentiality of study participants. The research team met regularly to resolve any discrepancies and to ensure consensus was reached.

*Ethics statement*

All participants provided written informed consent. Transcripts and survey responses were de-identified and stored in a secure and password-protected location that can only be accessed by members of the study team. Ethics approval was obtained from the UBC/Children's and Women's Health Centre of British Columbia Research Ethics Board (H23-03622).

**Reporting Checklists**

*Supplement 2: Consensus-Based Checklist for Reporting of Survey Studies (CROSS)*

| No. Item | Item Description | Reported on Page # |
|---|---|---|
| *Title and abstract* | | |
| 1a. Title and abstract | State the word "survey" along with a commonly used term in title or abstract to introduce the study's design. | P 2 |
| 1b. Title and abstract | Provide an informative summary in the abstract, covering background, objectives, methods, findings/results, interpretation/discussion, and conclusions. | P 2 |
| *Introduction* | | |
| 2. Background | Provide a background about the rationale of study, what has been previously done, and why this survey is needed. | Pp. 3-4 |
| 3. Purpose/aim | Identify specific purposes, aims, goals, or objectives of the study. | Pp. 3-4 |
| *Methods* | | |
| 4. Study design | Specify the study design in the "Methods" section with a commonly used term (e.g., cross-sectional or longitudinal) | P 4 and Suppl 1 |
| 5a. Data collection methods | Describe the questionnaire (e.g., number of sections, number of questions, number and names of instruments used). | P 5 and Suppl 1 |
| 5b. Data collection methods | Describe all questionnaire instruments that were used in the survey to measure particular concepts. Report target population, reported validity and reliability information, scoring/classification procedure, and reference links (if any). | P 5 and Suppl 1 |

| | | |
|---|---|---|
| 5c. Data collection methods | Provide information on pretesting of the questionnaire, if performed (in the article or in an online statement). Report the method of pretesting, number of times questionnaire was pretested, number of demographics of participants used for pretesting, and the level of similarity of demographics between pre-testing participants and sample population. | Suppl 1 |
| 5d. Data collection methods | Questionnaire, if possible, should be fully provided (in the article, or as appendices or as an online supplement). | P 5 and Suppl 5 |
| 6a. Sample characteristics | Describe the study population (i.e., background, locations, eligibility criteria for participant inclusion on survey, exclusion criteria) | P 4 and Suppl 1 |
| 6b. Sample characteristics | Describe the sampling techniques used (e.g., single stage or multiple stage sampling, simple random sampling, stratified sampling, cluster sampling, convenience sampling). Specify the locations of sample participants whenever clustered sampling was applied. | P 4 and Suppl 1 |
| 6c. Sample characteristics | Provide information on sample size, along with details of sample size calculation. | Suppl 1 |
| 6d. Sample characteristics | Describe how representative the sample is of the study population (or target population if possible), particularly for population-based surveys. | Suppl 1 |
| 7a. Survey administration | Provide information on modes of questionnaire administration, including the type and number of contacts, the location where the survey was conducted (e.g., outpatient room or by use of online tools, such as Survey Monkey). | P 5 and Suppl 1 |

| | | |
|---|---|---|
| 7b. Survey administration | Provide information of survey's time frame, such as periods of recruitment, exposure, and follow-up days. | Suppl 1 |
| 7c. Survey administration | Provide information on the entry process: For non-web-based surveys, provide approaches to minimize human error in data entry. For web-based surveys, provide approaches to prevent "multiple participation" of participants. | Suppl 1 |
| 8. Study preparation | Describe any preparation process before conducting the survey (e.g., interviewers' training process, advertising the survey). | Suppl 1 |
| 9a. Ethical considerations | Provide information on ethical approval for the survey if obtained, including informed consent, institutional review board [IRB] approval, Helsinki declaration, and good clinical practice [GCP] declaration (as appropriate). | P 5 and Suppl 1 |
| 9b. Ethical considerations | Provide information about survey anonymity and confidentiality and describe what mechanisms were used to protect unauthorized access. | Suppl 1 |
| 10a. Statistical analysis | Describe statistical methods and analytical approach. Report the statistical software that was used for data analysis. | N/A |
| 10b. Statistical analysis | Report any modification of variables used in the analysis, along with reference (if available). | N/A |
| 10c. Statistical analysis | Report details about how missing data was handled. Include rate of missing items, missing data mechanism (i.e., missing completely at random [MCAR], missing at random [MAR], or missing not at random [MNAR]), and methods used | N/A |

| | | |
|---|---|---|
| | to deal with missing data (e.g., multiple imputation). | |
| 10d. Statistical analysis | State how non-response error was addressed. | N/A |
| 10e. Statistical analysis | For longitudinal surveys, state how loss to follow-up was addressed. | N/A |
| 10f. Statistical analysis | Indicate whether any methods such as weighting of items or propensity scores have been used to adjust for non-representativeness of sample. | N/A |
| 10g. Statistical analysis | Describe any sensitivity analysis conducted. | N/A |
| *Results* | | |
| 11a. Respondent characteristics | Report numbers of individuals at each stage of the study. Consider using a flow diagram, if possible. | Fig 1 |
| 11b. Respondent characteristics | Provide reasons for non-participation at each stage, if possible. | P 6 and Suppl 1 |
| 11c. Respondent characteristics | Report response rate, present the definition of response rate or the formula used to calculate response rate. | Fig 1 |
| 11d. Respondent characteristics | Provide information to define how unique visitors are determined. Report number of unique visitors along with relevant proportions (e.g., view proportion, participation proportion, completion proportion) | Fig 1 and Suppl 1 |
| 12. Descriptive results | Provide characteristics of study participants, as well as information on potential confounders and assessed outcomes. | Pp. 5-6 and Suppl 1 |
| 13a. Main findings | Give unadjusted estimates and, if applicable, confounder-adjusted estimates | N/A |

| | | |
|---|---|---|
| | along with 95% confidence intervals and *p* values. | |
| 13b. Main findings | For multivariable analysis, provide information on the model building process, model fit statistics, and model assumptions (as appropriate), | N/A |
| 13c. Main findings | Provide details about any sensitivity analysis performed. If there are considerable amount of missing data, report sensitivity analyses comparing the results of complete cases with that of the imputed dataset (if possible). | N/A |
| *Discussion* | | |
| 14. Limitations | Discuss the limitations of the study, considering sources of potential biases and imprecisions, such as non-representativeness of sample, study design, important uncontrolled confounders. | P 11 |
| 15. Interpretations | Give a cautious overall interpretation of results, based on potential biases and imprecisions and suggest areas for future research. | Pp. 9-12 |
| 16. Generalizability | Discuss the external validity of the results. | Pp. 11-12 |
| *Other sections* | | |
| 17. Role of the funding source | State whether any funding organization has had any roles in the survey's design, implementation, and analysis. | Title page |
| 18. Conflict of interest | Declare any potential conflict of interest. | Title page |

| | | |
|---|---|---|
| 19. Acknowledgements | Provide names of organizations/persons that are acknowledged along with their contribution to the research. | Title page and P 13 |

*Supplement 3: Consolidated Criteria for Reporting Qualitative Studies (COREQ): 32-item checklist*

| No. Item | Guide questions/description | Reported on Page # |
|---|---|---|
| Domain 1: Research team and reflexivity | | |
| *Personal Characteristics* | | |
| 1. Interviewer/facilitator | Which author/s conducted the interview or focus group? | P 5 and Suppl 1 |
| 2. Credentials | What were the researcher’s credentials? E.g. PhD, MD | Suppl 1 |
| 3. Occupation | What was their occupation at the time of the study? | Suppl 1 |
| 4. Gender | Was the researcher male or female? | Suppl 1 |
| 5. Experience and training | What experience or training did the researcher have? | Suppl 1 |
| *Relationship with participants* | | |
| 6. Relationship established | Was a relationship established prior to study commencement? | Suppl 1 |
| 7. Participant knowledge of the interviewer | What did the participants know about the researcher? e.g. personal goals, reasons for doing the research | Suppl 1 |
| 8. Interviewer characteristics | What characteristics were reported about the interviewer/facilitator? e.g. Bias, assumptions, reasons and interests in the research topic | Suppl 1 |

| | | |
|---|---|---|
| Domain 2: study design | | |
| *Theoretical framework* | | |
| 9. Methodological orientation and Theory | What methodological orientation was stated to underpin the study? e.g. grounded theory, discourse analysis, ethnography, phenomenology, content analysis | P 5 and Suppl 1 |
| *Participant selection* | | |
| 10. Sampling | How were participants selected? e.g. purposive, convenience, consecutive, snowball | P 4 and Suppl 1 |
| 11. Method of approach | How were participants approached? e.g. face-to-face, telephone, mail, email | P 4 and Suppl 1 |
| 12. Sample size | How many participants were in the study? | Pp. 5-6, Fig 1, and Suppl 1 |
| 13. Non-participation | How many people refused to participate or dropped out? Reasons? | Fig 1 and Suppl 1 |
| *Setting* | | |
| 14. Setting of data collection | Where was the data collected? e.g. home, clinic, workplace | Suppl 1 |
| 15. Presence of non-participants | Was anyone else present besides the participants and researchers? | Suppl 1 |
| 16. Description of sample | What are the important characteristics of the sample? e.g. demographic data, date | Pp. 5-6 and Suppl 1 |
| *Data collection* | | |
| 17. Interview guide | Were questions, prompts, guides provided by the authors? Was it pilot tested? | Yes, see Suppl 1 and 5 |
| 18. Repeat interviews | Were repeat interviews carried out? If yes, how many? | Suppl 1 |

| | | |
|---|---|---|
| 19. Audio/visual recording | Did the research use audio or visual recording to collect the data? | P 5 and Suppl 1 |
| 20. Field notes | Were field notes made during and/or after the interview or focus group? | Suppl 1 |
| 21. Duration | What was the duration of the interviews or focus group? | Suppl 1 |
| 22. Data saturation | Was data saturation discussed? | Suppl 1 |
| 23. Transcripts returned | Were transcripts returned to participants for comment and/or correction? | Suppl 1 |
| Domain 3: analysis and findings | | |
| *Data analysis* | | |
| 24. Number of data coders | How many data coders coded the data? | P 5 and Suppl 1 |
| 25. Description of the coding tree | Did authors provide a description of the coding tree? | P 6 and Tables 2-6 |
| 26. Derivation of themes | Were themes identified in advance or derived from the data? | P 5 and Suppl 1 |
| 27. Software | What software, if applicable, was used to manage the data? | P 5 and Suppl 1 |
| 28. Participant checking | Did participants provide feedback on the findings? | Pg 5 and Suppl 1 |
| *Reporting* | | |
| 29. Quotations presented | Were participant quotations presented to illustrate the themes/findings? Was each quotation identified? e.g. participant number | Yes, see pp. 6-9 and Tables 2-6 |

|  |  |  |
|---|---|---|
| 30. Data and findings consistent | Was there consistency between the data presented and the findings? | Yes, see pp. 6-9 and Tables 2-6 |
| 31. Clarity of major themes | Were major themes clearly presented in the findings? | Yes, see pp. 6-9 and Tables 2-6 |
| 32. Clarity of minor themes | Is there a description of diverse cases or discussion of minor themes? | Yes, see pp. 6-9 and Tables 2-6 |

*Supplement 4: Guidance for Reporting Involvement of Patients and the Public (GRIPP2) short form*

| Selection and topic | Item | Page # |
|---|---|---|
| 1: Aim | *Report on aim of PPI in the study*<br><br>Despite the well-documented benefits of patient engagement and increasing prevalence of patient engagement in sepsis research and advocacy, there is limited understanding of the impacts of this involvement on sepsis survivors and their families. The primary aim of this PPI was to investigate the long-term impacts of involvement in patient advisory councils.<br><br>A secondary aim of PPI in the study was to support sepsis research that addressed the priorities of sepsis survivors and their families. The research question and study design were co-developed with members of the Patient Advisory Councils (PACs) of two sepsis research networks, and additional members were co-investigators for the study. These individuals either participated in or were consulted during data analysis and interpretation to ensure findings were aligned with patient perspectives. | Pp. 1-4 and Suppl 1 |
| 2: Methods | *Provide a clear description of the methods used for PPI in the study*<br><br>This mixed-methods study was initiated and led by two members of Sepsis Canada and Action on Sepsis PACs and a UBC Assistant Professor (all co-PIs). The study team also included the Action on Sepsis network coordinator and a PhD candidate serving on the Sepsis Canada Steering Committee. We engaged | Pp. 4-5 and Suppl 1 |

| | | |
|---|---|---|
| | additional patient partners (n=4) to inform the development of data collection tools and conduct data analysis.<br><br>We first conducted a quantitative survey using the Patient Engagement In Research Scale. Patient partners (n=4, including the co-PIs) participated in adapting the survey tool and interpreting the results, which were used to inform the development of our interview guide. All current members (n=29) of the Sepsis Canada and Action on Sepsis PACs were invited to participate. The gender (primarily women), age (20-80 years), and geographic distribution (residents of 6 provinces) of participants were representative of the existing PACs, which includes both sepsis survivors and caregivers of individuals who experienced sepsis. A research assistant who had not previously engaged with any members of the PACs conducted 10 independent interviews and 1 focus group to gain an in-depth understanding of patient partners' experiences. Three members of the research team reviewed the transcripts of three interviews and developed an initial coding framework. This framework was discussed with a patient partner until all coding frameworks were agreed. Coded transcripts were de-identified and shared with two patient partners and a researcher. The interpretation of data was discussed regularly until a consensus on coding was reached. | |
| 3: Study results | *Outcomes – Report the results of PPI in the study, including both positive and negative outcomes.*<br><br>Patient partners contributed to the study in the following ways: initial conceptualization of the study; ensuring that the survey, interview guide, and other materials were trauma-informed; co-developing coding framework and reviewing coded transcripts; analyzing the results through the lens of their sepsis experiences; and reviewing the manuscript to ensure that the study objectives and findings were reflective of their priorities. Additionally, the research assistant and patient partner co-presented a poster on this study at a conference focused on patient-oriented research in BC. | Pp. 6-9 and Tables 2-6 |

| | | |
|---|---|---|
| 4. Discussion and conclusions | *Outcomes – Comment on the extent to which PPI influenced the study overall. Describe positive and negative effects.*<br><br>PPI was critical in the study as it informed all aspects of the study design. Their involvement ensured the research question remained relevant to patient partners and that potential risks to patient partners who were participants in the study were properly mitigated, as reflecting on sepsis experiences can be triggering for survivors and their family members. Patient partners had prior experience with being involved in research as study participants and as peer researchers, which allowed for a unique perspective to be included in this study, and the study team was able to support patient partners in building additional research capacity that complement their existing training and involvement with the sepsis research networks.<br><br>Engagement with PACs members through this study also allowed for the researchers to better understand the importance of patient engagement, and patient priorities and expectations in their engagement. This will allow for better partnership in future studies. | Suppl 1 |
| 5. Reflections/critical perspective | *Comment critically on the study, reflecting on the things that went well and those that did not, so others can learn from this experience.*<br><br>Patient partners in the study collaborated well and were excited to see their research question come to life. We were able to reflect the study findings to our own engagement with PACs members which enhanced the quality and satisfaction of this partnership. However, a challenge was communication and meeting project timelines, as patient partners had additional responsibilities and roles beyond the study. This is manageable but needs to be recognized in the planning stage. Ensuring there is a study team member who can provide dedicated logistical support and study coordination can also help with managing this challenge. | Suppl 1 |

PPI = patient and public involvement

**Supplement 5: Survey**

**Section 1: Experience with Sepsis**

1. Which of the following best described your experience with sepsis? Please select all that apply. (Options: Sepsis survivor, family member or caregiver of individual who experienced sepsis)
2. How long ago was your most recent sepsis experience? (e.g., 2 years and 3 months):
   Years: ____ Months: ____
3. How many different encounters have you had with sepsis?
4. How long ago was your first sepsis experience? (e.g., 2 years and 3 months ago):
   Years: ____ Months: ____

**Section 2: Experience within the Patient Advisory Councils**

5. Which research network do you belong to? (Options: Sepsis Canada, Action on Sepsis)
6. How long have you been a member? (e.g., 2 years and 3 months):
   Years: ____ Months: ____
7. What type of projects have you supported in your role with the network? Please select all that apply.

(Options:

- Network Operations (e.g., Participating in a strategic planning workshop, Attending a steering committee meeting or Patient Advisory Council Meeting);
- Research Activities (e.g., Reviewing a grant proposal or manuscript, Attending lab/team meetings to learn about or inform sepsis research, Co-developing and or leading a research study with network members);
- Knowledge translation (e.g., Informing, participating in or leading sepsis public awareness campaigns; Speaking to students, health workers, researchers, or the public at educational symposia or lectures; Supporting or informing educational materials for patients on sepsis or post-sepsis care).
- Other (open-text response))

8. Since joining the Patient Advisory Council, on average how many hours per month have you spent on work related to your role on the Council?
9. Did you have prior experience serving as a patient partner for a research network or research project? (Options: Yes, No)

For the next series of questions, please think about your experience as a patient partner with Sepsis Canada or Action on Sepsis and respond to the statements by ticking only one box for each statement. If you are unsure about which option to choose for a statement, please give the best response you can.

For the purposes of these questions, a project can be any of the activities you have supported in your role with either research network. If you have taken part in multiple projects, please select the response that best captures your overall experience.

All questions have following response options:

| Strongly Agree | Agree | Neutral | Disagree | Strongly Disagree | Not Applicable |
|---|---|---|---|---|---|
| ☐ | ☐ | ☐ | ☐ | ☐ | ☐ |

### Procedural Requirements

The following fourteen (14) statements are about your general experiences throughout the project:

PR1. I was interested in the issue(s) being researched in the project
PR2. The research team members were properly introduced to each other
PR3. The number of patient partners on the research project team seemed appropriate
PR4. I understood the objective(s) of the project
PR5. I agreed with the objective(s) of the project
PR6. I understood how I could contribute to the project
PR7. I received sufficient explanation about the project
PR8. I understood my ethical responsibilities for the project
PR9. In general, I had sufficient opportunities to contribute to the project
PR10. I was able to perform my tasks for the project
PR11. I participated in making decisions about the project
PR12. I received sufficient updates about the project
PR13. Communication within the research team was clear throughout the project
PR14. The project was worth the time I spent on it

### Convenience

The following four (4) statements are about how convenient it was for you to contribute throughout the project:

CN1. I had the opportunity to provide input into selecting my tasks for the project
CN2. My preferences for meetings (such as time, duration, location, and format) were considered when planning meetings
CN3. Throughout the project, I had sufficient time to complete my tasks for the project
CN4. I had opportunities to express my views

### Contributions

The following four (4) statements are about your contributions throughout the project:

CT1. I contributed by providing my perspective
CT2. My contributions were a good use of my time
CT3. I shared my knowledge within the project team
CT4. My workload in the project was manageable

### Team Environment and Interaction

The following five (5) statements are about the research environment and interaction throughout the project:

T1. Throughout the project, I felt accepted as a member of the research project team
T2. I was an equal partner in the research project team
T3. My interactions within the research project team were positive
T4. There was mutual respect among the research project team members

T5. There was trust among the research project team members

Support

The following three (3) statements are about the support provided throughout the project:

SU1. I received sufficient support to contribute to the project (for example, orientation, readings, training workshops, webinars)

SU2. Any concerns I had were addressed

SU3. I was offered sufficient reimbursement for my out-of-pocket expenses (such as childcare, parking, and travel) related to the project activities

Feel Valued

The following three (3) statements are about your feeling of being a valued member of the research team.

FV1. The research project team appreciated my contributions

FV2. The research project team was open to receiving my views

FV3. I was offered sufficient recognition for my contributions (for example, payment, authorship, or gifts)

Benefits

The following four (4) statements are about the benefits of your involvement in the project:

BE1. I enjoyed being a part of the project

BE2. I made an impact on the decisions in the project

BE3. I saw how my contributions could benefit others

BE4. My involvement had positive impacts on my life

Is there anything else you wish to share about your role and experiences as a member of a sepsis research network, or the impact it has had on your sepsis or post-sepsis journey? (Open-ended response)

**Section 3: Demographics**

10. Where do you currently reside? (Options: Alberta, British Columbia, Manitoba, New Brunswick, Newfoundland and Labrador, Nova Scotia, Ontario, Prince Edward Island, Quebec, Saskatchewan, Northwest Territories, Nunavut, Yukon, I prefer not to answer)

11. Which age group do you belong to? (Options: <20 years, 20-30 years, 30-40 years, 40-50 years, 50-60 years, 60-70 years, 70-80 years, 80-90 years, > 90 years, I prefer not to answer)

12. What is your gender? (Options: Woman, Man, Non-binary, Genderfluid or genderqueer, I do not know, I prefer not to answer, Another gender identity (please specify): open-text)

13. What is the highest level of education you have completed? (Options: Some high school, no diploma; High school graduate, diploma or the equivalent (for example: GED); Postsecondary (for example: Trade/technical/vocational training, Bachelors); Postgraduate (for example: Masters or Doctorate); I prefer not to answer; Other (open-text))

14. What was your employment status before your sepsis experience? (Options: Full-time employee, Part-time employee, Self-employed, Stay-at-home parent/caregiver, Retired, Unemployed, I prefer not to answer, Other (open-text))
15. Have you returned to the same employment status following your sepsis experience? (Options: Yes, No, I prefer not to answer)
16. How many months did it take you to return to the same employment status? Leave this blank if you can't remember.

**Supplement 6: Interview Guide**

*Introduction*

Which network are you part of? What drew you to the council/network?

*Sepsis Recovery & Impact of Engagement*

1) Can you tell me about your sepsis recovery journey? What did you expect your recovery to look like?

Probes: Did you expect to be able to return to work shortly after? What impacts did you expect it to have on your physical health? Mental health? Social network? How long did you expect these impacts to take?

2) Did your participation in the council impact your recovery in any way? In what way?

Probes: How did your recovery change over the course of your involvement with the council? Were there different impacts on your physical recovery? Mental recovery? Social support system? Were there factors in the way the council operates that impeded or supported your recovery?

3) Can you tell me about your experience participating in the patient council?

Probes: What types of activities did you participate in; how long have you been involved in the council, how much time do you commit on the average week/month/year? How frequent are the related activities and meetings? How was the council organized? What type of support were you provided with? What type of recognition/compensation do you receive?

*Experience with the Council*

4) Can you tell me about your expectations for involvement in the council? What did you expect your participation would look like before you started? Did it meet these expectations - how or how not?

Probe: Did you have a role in decision-making? Did you expect more or less support? Did you expect more or less opportunities to share your perspective? What knowledge or information did you contribute?

5) What motivated you to join the council? Do you feel your contributions are valued?

Probes: What made you feel this way? Did this change over time or depend on the specific activity you were involved in? Did this change depending on the people who worked with?

6) What made participating in the council easy and/or challenging?

Probes: Did the meeting times interfere with other commitments or time constraints? Did you have input into when meetings occurred? Did other team members provide support? Were you provided with resources or training to support your participation? Did the format of the meeting include opportunities to contribute?

7) Did you form any new personal or professional relationships as a result of your involvement in the council?

If yes - Did these relationships extend to activities outside of those organized or supported by the council? What impact did these relationships have on your recovery from sepsis?

8) Do you think your participation in the council impacted others' recovery from sepsis?

Probes: For example, what about your family members? Other council members? General public?

9) What impact do you think your involvement with the council will have on your recovery in the future?

Probes: Do you think it will impede your recovery, support it, or have no effect? Why or why not?

10) Would you recommend other sepsis survivors/family members of sepsis survivors participate in patient advisory councils? Why/why not?

Probes: Are the activities or time and energy commitment appropriate? Would they agree with the objectives? Do you think they could benefit from the experience? Has the experience with other members been positive or negative?

*Closing*

11) If you could make changes to the way the council operates, what would you recommend?

Probes: activities, objectives, time/energy commitment, processes for recruiting or engaging with members, support provided, recognition provided

12) How would you recommend we share lessons learned from this study with yourself and other patient partners in the future? Probe: Format (i.e., presentation vs online resource vs brief report)
